# Detection of local microvascular proliferation in *IDH* wild-type Glioblastoma using relative Cerebral Blood Volume

**DOI:** 10.1101/2021.04.19.21255589

**Authors:** María del Mar Álvarez-Torres, Elies Fuster-García, Javier Juan-Albarracín, Gaspar Reynés, Fernando Aparici-Robles, Jaime Ferrer-Lozano, García-Gómez Juan Miguel

**Affiliations:** Universitat Politècnica de València, ITACA. Valencia, Valencia, Spain. 46022; Oslo University Hospital, Department of Diagnostic Physics. Oslo, Norway. 0424; Health Research Institute Hospital La Fe, Department of Medical Oncology, Cancer Research Group. Valencia, Valencia, Spain. 46026; Health Research Institute Hospital La Fe, Department of Medical Imaging. Valencia, Valencia, Spain. 46026; Health Research Institute Hospital La Fe, Department of Pathology. Valencia, Valencia, Spain. 46026

**Keywords:** *IDH* wild-type glioblastoma, microvascular proliferation, cerebral blood volume, perfusion imaging, microvessel area

## Abstract

**Background:** The microvascular proliferation (MVP) and the microvessel area (MVA) are known as diagnostic and prognostic biomarkers for glioblastoma; nevertheless, its measurement is costly, labor-intense, and invasive. MRI perfusion biomarkers such as such as relative cerebral blood volume (rCBV) may be a feasible alternative to predict MVP and estimate MVA.

**Purpose:** This study aims to evaluate the detection capacity of MRI markers such as rCBV to detect local microvascular proliferation in IDH wild-type glioblastoma. In addition, we aim to analyze the association between rCBV values and the microvessel area in different regions of the tumor.

**Study type:** Retrospective study.

**Population and subjects:** Data from 71 tissue blocks belonging to 17 *IDH* wild-type glioblastoma patients were compiled from the Ivy GAP database.

**Field Strength/Sequence:** 1.5T or 3.0T. Pregadolinium and postgadolinium-based contrast agent-enhanced T1-weighted MRI, T2- and FLAIR T2-weighted, and dynamic susceptibility contrast (DSC) T2* perfusion.

**Assessment:** We analyzed preoperative MRIs to establish the association between the maximum and mean relative cerebral blood volume (rCBV_max_ and rCBV_mean_) with the presence/absence of microvascular proliferation and with the microvessel area for each tumor block.

**Statistical tests:** Spearman’s correlation and Mann-Whitney test.

**Results:** Significant positive correlations were found between rCBV and MVA in the analyzed tumor blocks (p<0.001). Additionally, significant differences in rCBV were found between blocks with MVP and blocks without MVP (p<0.0001).

**Data conclusion:** The rCBV is shown as significantly different in those tissue blocks with microvascular proliferation from those blocks without it, and it is significantly correlated with microvessels area. This method allows a local detection and definition of MVP and MVA in different regions of the glioblastoma since the first diagnostic stage and in a non-invasive way.

## INTRODUCTION

*IDH* wild-type glioblastoma is the most lethal and common tumor of the central nervous system [1, 2], characterized by high vascularity [3-5]. Blood supply is required for the establishment, growth, and progression of the tumor; and several mechanisms are implicated in the formation of new vessels: hypoxia/angiogenesis from pre-existing vessels, secretion of angiogenic factors that recruit cells for new vessel formation (vasculogenesis), and incorporation of tumor cells into the vascular endothelium [3-5]. One of the results of these mechanisms is microvascular proliferation (MVP), which generally occurs in the core of glioblastomas by sprouting new vessels from pre-existing ones, depending on the presence of hypoxia [3].

MVP is marked by two or more blood vessels sharing a common vessel wall of endothelial and smooth muscle cells [5], and interactions between tumor cells and blood vessels during microvascular proliferation seem to facilitate tumor growth [5-7]. The result of microvascular proliferation is the formation of large-lumen microvessels, usually with a glomeruloid appearance, that represent a histopathologic hallmark of glioblastoma [8].

Considering the relevance of this vascular process, related histological features, such as the microvessel area (MVA), i.e., the total area covered by the microvessels in the tumor sample, and microvessel density (MVD), i.e., the number of microvessels per volume unit, have been previously investigated [8-10, 16-20]. Different studies suggest that MVD poorly describes the morphometric diversity of these microvessels in high-grade gliomas [8-10]. However, MVA may provide a more robust clinical biomarker, useful for prognosis and grading [8-15], since it has been shown as correlated with patient survival [8-12] and also with higher histological grade of gliomas [8, 13, 14, 15].

Regardless of this evidence, the histopathological quantification of MVA is still used exclusively in the research setting. Relevant limitations, including time- and cost-expending, labor intensity, and invasiveness make it challenging for routine clinical practice.

An alternative approach previously studied in the literature to overcome the limitations in MVA quantification through histopathological analysis is perfusion MRI [8, 16]. Some studies found that measures of relative cerebral blood volume (rCBV) positively correlate with microvascular structures in different glioma tumors [8, 10, 17-20]. However, these studies are few and present important limitations such as animal-based studies [10, 17], small cohorts of glioblastoma patients [8, 18-20], low number of analyzed histopathological specimens [8, 18-20], or analysis with non-spatial coregistered data [14, 17].

Glioblastoma presents higher MVA than low-grade tumors, healthy brain, and non-tumoral processes like inflammation and post-treatment effect [16]. This basic hallmark underlies different noteworthy clinical applications, including predicting prognosis, grading, quantification of tumor cell density and extent, characterization of molecular/genomic profiles, and discrimination between tumor progression from non-tumoral post-treatment effects [16].

Therefore, the integration of advanced and automatic techniques capable of calculating robust imaging parameters, like rCBV, could help address many unmet goals in the glioblastoma patient’s management. Besides, the use of rCBV would involve important advantages since calculations derived from routine MRIs can be performed through automatic and robust methodologies [21-23] and in a non-invasive way.

In this context, considering that MVA is related to the microvascular proliferation process, and a significant correlation has been found between perfusion MRI and glioma vascular features in previous studies, we hypothesize that MVA could be directly associated with the rCBV in *IDH* wild-type glioblastoma and this correlation can be measured using a robust MRI processing service. The areal density of microvessels on sections is an unbiased estimator of the volume density of microvessels according to the Delesse principle [24], and we hypothesize that the volume of microvessels can be related to the rCBV. Since the typical spatial resolution, DSC sequences is 2-mm in-plane × 5-mm slices [25], the calculation of rCBV would be reliable when it is calculated in areas larger than 2mm.

The general purpose of our study is to evaluate the potential use of rCBV, calculated with the ONCOhabitats platform, to detect microvascular proliferation in different regions of *IDH* wild-type glioblastoma, using the initial standard MRI studies. The limitations of the previous studies (animal based-studies [10, 17], small cohorts [8, 18-20], few tissue samples [8, 18-20], non-registered data [14, 17], or non-continuous analyzed variables [14, 17, 20]) have been tried to overcome in this study. All data was collected from the public database Ivy Glioblastoma Atlas Project (Ivy GAP), and rCBV maps were calculated using the public services included in the ONCOhabitats website (https://www.oncohabitats.upv.es), to ensure the reproducibility and repeatability of the study. The study’s specific objectives are 1) to analyze the correlation between the imaging markers (rCBV_mean_ and rCBV_max_) with the MVA of different glioblastoma regions and 2) to study whether these imaging markers can differentiate regions of the tumor that present MVP.

## MATERIALS AND METHODS

### Clinical data collection

The Ivy Glioblastoma Atlas Project (Ivy GAP) database (www.glioblastoma.alleninstitute.org) [26] was used for this study. This public database includes 41 verified glioblastoma patients, with a total of 42 tumors, with the following information per tumor: 1) images of the resected tumor (Figure 1A.I) divided into tissue blocks (Figure 1A.II); 2) histopathological images at a cellular resolution of hematoxylin and eosin-stained sections (collected from the tissue blocks) annotated for anatomic structures, including areas of MVP (Figure 1A.III and Figure 1B), and 3) pre-surgical MRI studies of the patients: including pre and post-gadolinium T1-weighted MRI, T2-weighted MRI, FLAIR and Dynamic Susceptibility Contrast (DSC) T2* perfusion-weighted MRI (Figure 2A).

**Figure 1.**
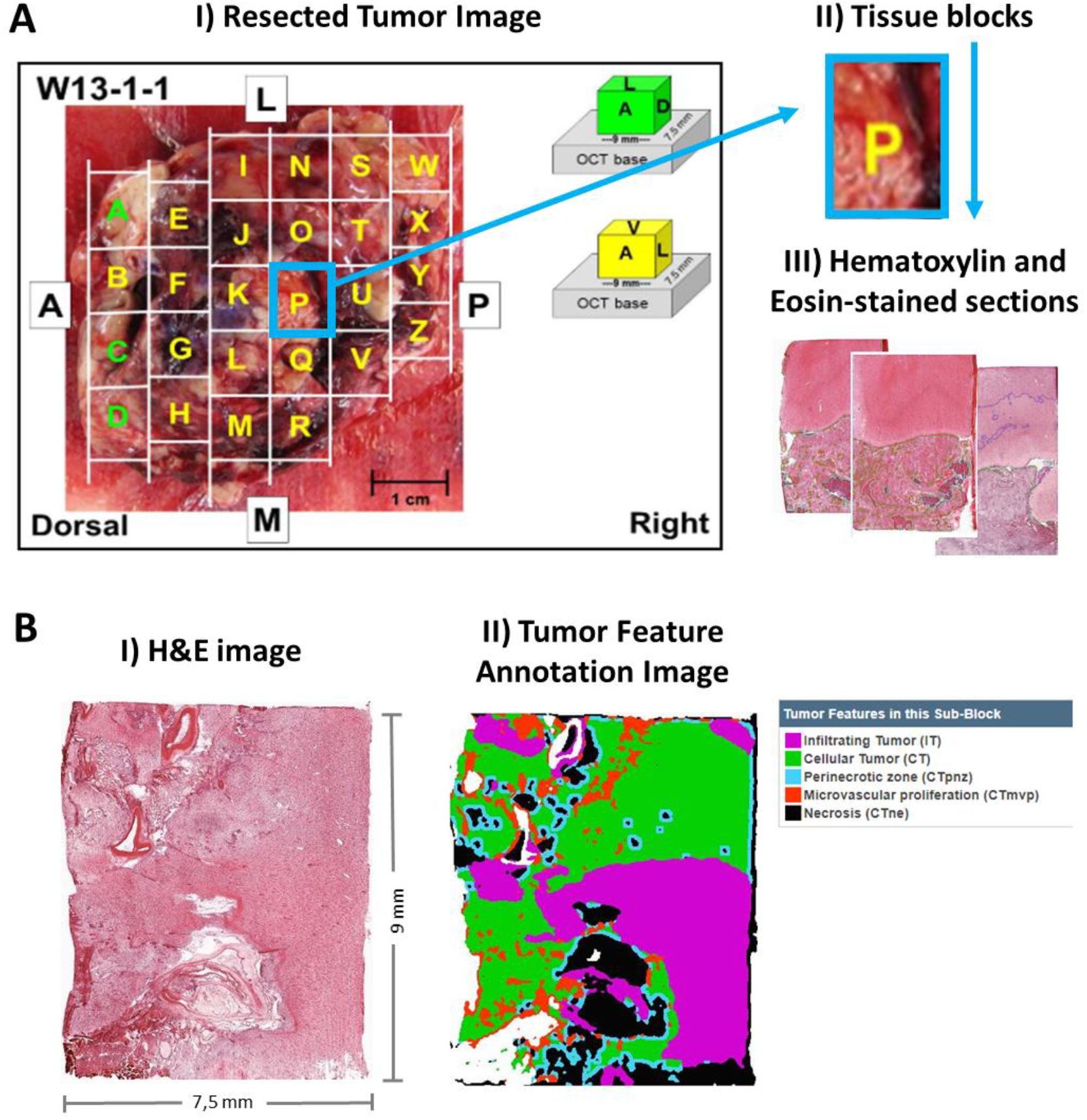
**A**: Included information in the Ivy Glioblastoma Atlas Project (Ivy GAP) database: I) Resected Tumor Image II) divided in tissue blocks; and III) Histopathological images at cellular resolution of hematoxylin and eosin-stained sections. **B**: I) Example of an H&E image of a slide from a resected block. II) The same image labeled with the different tissues and structures. Microvascular proliferation corresponds with areas in red color. (Images from Ivy GAP database [26], patient W55, block F, slice F.02).

**Figure 2.**
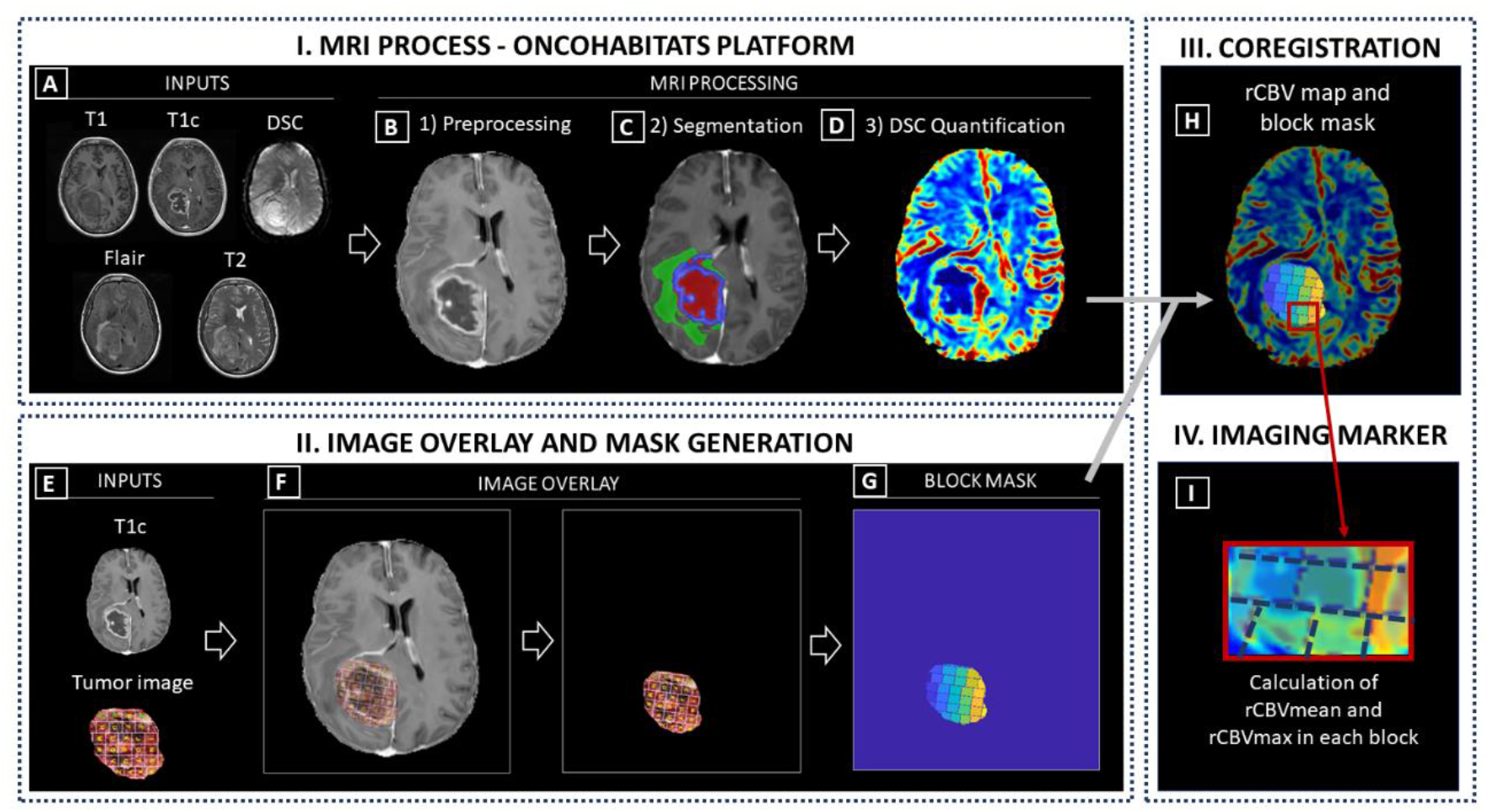
Phases of methodology: I) MRI Process conducted by the Segmentation Service included in the ONCOhabitats platform; II) Image overlay and mask generation; III) Coregistration and IV) Calculation of imaging markers. (A) Required inputs for the MRI process (T1, T1c, T2, Flair and DSC); (B) Preprocessing of the MRIs; (C) Classic segmentation of the lesion into tumor, edema and necrosis; (D) DSC perfusion quantification and rCBV map calculation. (E) Inputs to generate the block mask; (F) Image overlay process for mask generation; (G) Block mask; (H) Overlay of the rCBV map and block mask; (I) Calculation of imaging markers (rCBV_mean_ and rCBV_max_) in each tissue block.

### Tumor selection

Inclusion criteria for patients participating in the study were: *i)* histopathological confirmation of glioblastoma *IDH* wildtype; *ii)* access to complete MRI studies, including pre and post-gadolinium T1-weighted (T1 and T1c, respectively), T2-weighted, FLAIR T2-weighted, and DSC T2* perfusion sequences (Figure 2A); *iii)* access to the resected tumor image (Figure 1A); and *iv)* approval by an expert radiologist and histopathologist of the correct overlay/registration of the image of the resected tumor and the MRI of each patient (Figure 2)

### MRI acquisition and processing

MR images were obtained on a 1.5 or 3 T scanner (Siemens Healthcare, Erlangen, Germany or GE Medical Systems, Waukesha, Wisconsin) [26]. MRI processing was carried out using the ONCOhabitats platform [23], freely available in https://www.oncohabitats.upv.es. The ONCOhabitats analysis included the following automatic stages (Figure 2):

1. ***MRI Preprocessing*** (Figure 2B), including voxel isotropic resampling of all MR images, correction of the magnetic field in homogeneities and noise, rigid intra-patient MRI registration, and skull-stripping (Figure 2A).
2. ***Glioblastoma tissue segmentation*** (Figure 2C), performed by using an unsupervised segmentation method, which implements a state-of-the-art deep-learning 3D convolutional neural network (CNN), which takes as input the T1c, T2, and Flair MRI. This method is based on Directional Class Adaptive Spatially Varying Finite Mixture Model, or DCA-SVFMM, which consists of a clustering algorithm that combines Gaussian mixture modeling with continuous Markov random fields to take advantage of the self-similarity and local redundancy of the images. The result is the automated delineation of the enhancing tumor, edema, and necrosis regions with an overall performance of 0.9 Dice in whole tumor segmentation.
3. ***DSC perfusion quantification*** (Figure 2D), which calculates the relative cerebral blood volume (rCBV) maps, as well as relative cerebral blood flow (rCBF) or Mean Transit Time (MTT), for each patient. In this phase, T1-weighted leakage effects are automatically corrected using the Boxerman method [27], while gamma-variate curve fitting is employed to correct for T2 extravasation phase. rCBV maps are calculated by numerical integration of the area under the gamma-variate curve. The Arterial Input Function (AIF) is automatically quantified with a divide and conquer algorithm.

A more detailed description of the methodology is included in Reference 23, and the results of a multicenter study demonstrated its robustness in included in Reference 21.

### ISH Image Acquisition and Processing

Whole slides were scanned directly to SVS file format at a resolution of 0.5μm/pixel without downsampling on ScanScope® scanners (Aperio Technologies, Inc; Vista, CA) equipped with a 20x objective and Spectrum software. The raw image files of ∼5 GB per image were archived after converting to JPEG 2000 file format. The preprocessed images were flipped along the horizontal axis, white balanced, and compressed at a rate of 0.8 to ∼400 MB per image. During post-processing, colorized expression values or heat masks showing ISH signal intensity were generated, and the closest H&E stained image of the same specimen was calculated for each ISH section.

During the review of images, the automated bounding box overlay was manually adjusted if necessary, so that each of 8 bounding boxes per slide was placed over the corresponding tissue section, and images of slides with focus or image tile stich misalignments were re-scanned. Images were failed if artifacts compromised data analysis (e.g., mechanical damage, mounting medium bubbles, hybridization bubbles, and NBT/BCIP precipitated aggregates) associated with the corresponding tissue section.

### Image overlay, mask generation, and image markers

To compare the information obtained from the rCBV maps and the MVA obtained from histopathological images, we overlaid the T1c MRI images to the images of tumor resected pieces, including the histopathology blocks’ location (Figure 2E) provided in the Ivy GAP dataset. This 2D registration was performed manually considering the size, orientation, and location of the tumor, and expert radiologists indications (Figure 2F). Additionally, we remove the background and generate a block mask with each block area delimited (Figure 2G).

Once the blocks were coregistered with the MRI space (2H), we could obtain the imaging markers rCBV_mean_ and rCBV_max_ for each independent block (Figure 2I).

### Study variables

From each tissue block, several slides with an area of approximately 9 × 7,5mm were collected with their corresponding hematoxylin and eosin (H&E) images of the IVY Gap database. The areas of different histopathological tissues were delimited in these images (Figure 1B) and quantified data was available, including the mean value of MVA per block and the total area of each section of the block.

In order to normalize the MVA according to the area of each section of the tumor, the MVA value was divided by the area of the section. In addition, each block contains information from different sections; therefore, for the statistical analyses, we used the normalized mean value of MVA for each block, calculated with the following formula (where MVA_bs_ is the MVA present in the section s of one particular block b where n sections are available):

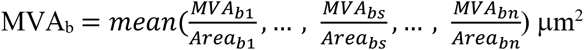

Besides, each block was classified into two groups: the *microvascular proliferation group (MVP group)* and the *non-microvascular proliferation group (non-MVP)*, depending on the presence (MVA>0) or absence of microvessels (MVA=0). An example of the vessels generated by microvascular proliferation derived from the tumor progression, as opposed to normal vessels, is illustrated in Figure 3.

**Figure 3.**
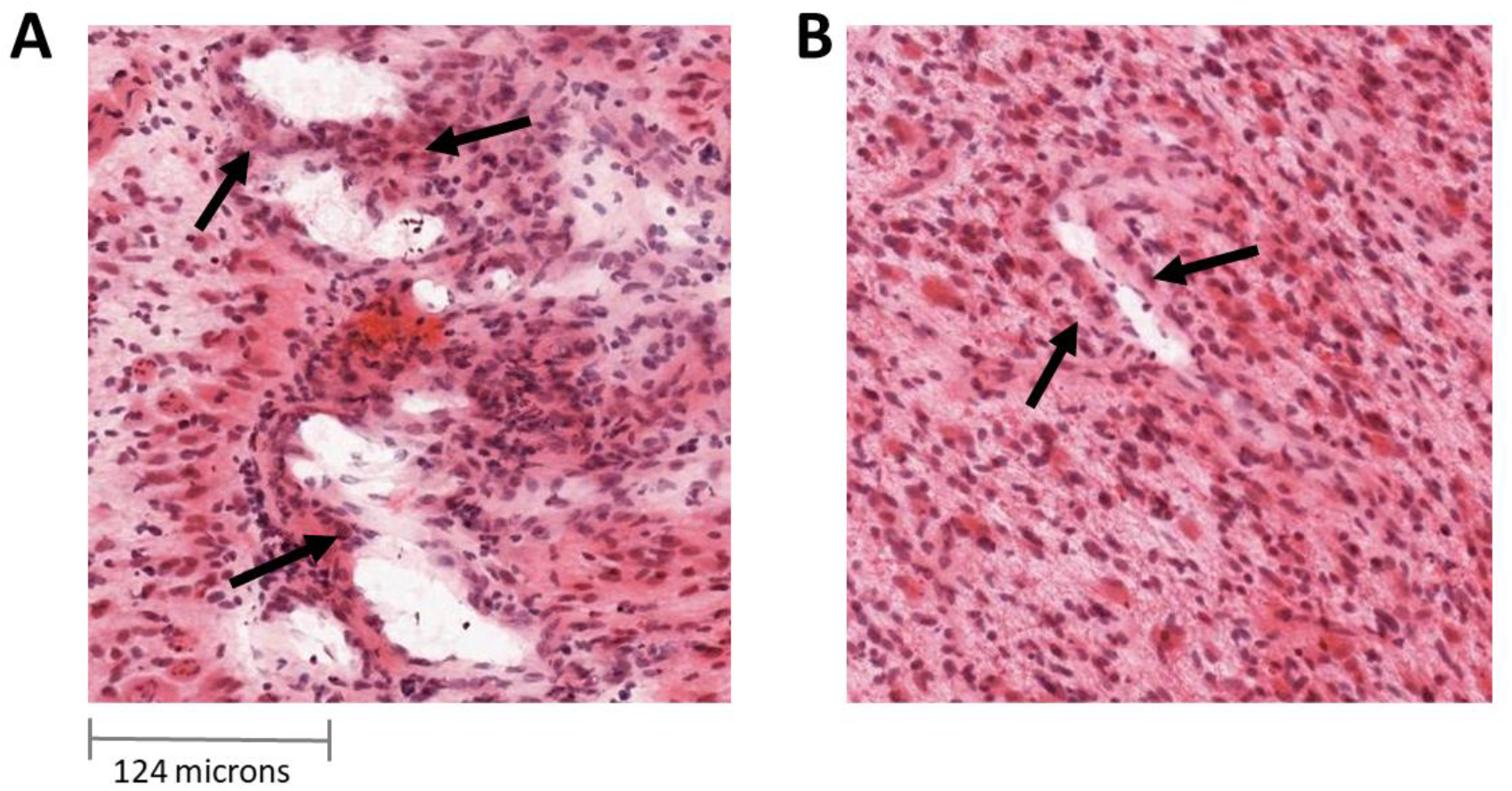
**A**: Aberrant vessels generated by microvascular proliferation sharing common vessel walls of endothelial and smooth muscle cells. **B**: Normal vessels. (Images from Ivy GAP database).

### Comparison with Previous Studies and Statistical Analyses

This study comprises three main analyses: (1) to compare the main characteristics of this study with those of previously published studies, (2) to assess the association between maximum rCBV (rCBV_max_) and mean rCBV (rCBV_mean_) and microvessel area, and (3) to analyze whether significant differences exist in rCBV_max_ and rCBV_mean_ between *MVP group* and *non-MVP group* in different regions of the tumors. Statistical analyses were performed with MatLab R2017b (MathWorks, Natick, MA).

#### Comparison with previous studies

A comparative analysis was carried out to compare the previously published studies with this study. The following relevant variables were analyzed:

- Nature of the study cohort: human or animal specimens (Human: Yes/No).
- The number of included individuals with glioblastoma WHO grade IV (or other glioma types) (#Patients).
- The number of analyzed tissue samples (#Tissue samples).
- Use of the standard initial MRI, routinely used for tumor diagnosis (Yes/No).
- Spatial coregistration between tissue samples and MRI (Yes/No).
- Analysis of continuous histopathological variables (area or volume of microvascular structures) (Yes/No).

#### Association between rCBV and MVA

Spearman’s correlation test was performed to study the association between the rCBV_mean_ and rCBV_max_ with the microvascular proliferation area (numeric continuous variable). Spearman coefficients and derived p-values were calculated, and descriptive measures of both rCBV_mean_ and rCBV_max_ among the whole block samples (mean, standard deviation, median, and range).

#### Differences of rCBV between MVP group and Non-MVP group

Mann-Whitney tests were conducted to analyze the differences of rCBV_mean_ between the *MVP group* and the *non-MVP group*. The descriptive measures of rCBV (mean, standard deviation, and range) for each MVP and non-MVP groups were included.

## RESULTS

### Included Tumors

From the 41 patients included in the Ivy GAP database, 3 were discarded because of non-histopathological confirmation of *IDH* wild-type glioblastoma (W10, W31, and W35); 14 were not included because of incomplete MRI studies (W04, W06, W16, W19, W20, W21, W26, W27, W28, W32, W39, W45, W53, and W54); 3 patients were discarded due to inability to correctly overlay the image of the resected tumor over the MRI (W08, W09, and W11). The remaining 21 patients were processed with the ONCOhabitats platform.

Of these 21 cases, 3 were excluded due to glioblastoma tissue segmentation errors (W01, W03, and W22); and 1 patient was excluded because of extensive hemorrhage that prevented a correct quantification of perfusion maps in DSC-MRI (W50). Finally, 17 tumors were included in the study.

### Included Blocks

A total of 124 blocks with complete information from the 17 tumors was initially considered for the study. However, to develop the statistical analyses, we selected all those blocks with more than twenty-five percent (> 25%) of tumor tissue, according to the imaging segmentation. This criterion was based on the existence of blocks that mostly contain necrotic tissue and therefore were not suitable for the study due to their lack of vascularization. 71 tissue blocks formed the final study sample.

### Comparison with previous studies

After searching the main scientific databases (Google Scholar, Pubmed, and the Web of Science), we found seven published articles related to the correlation between rCBV and vascular variables measured by histopathology [8, 10, 17-21]. Table 1 includes the comparison of the main characteristics of each study.

**Table 1:** Comparative table with previous studies reported in literature related with the correlation between MRI and vascular features defined by histopathological analyses.

|  | Human cohort | #Patients | #Tissue samples | Diagnostic MRI<sup>a</sup> | Spatial coregistration<sup>b</sup> | Continuous histopathological variable<sup>c</sup> |
| --- | --- | --- | --- | --- | --- | --- |
| Aronen HJ *et al.*; 1994 [14] | ✓ | 5 WHO IV (+14 other gliomas) | Not specified | ✗ | ✗ | ✗ |
| Cha S *et al.*; 2003 [17] | ✗ | 34 mice | 34 | NA | ✗ | ✗ |
| Chakhoyan A *et al.*; 2019 [18] | ✓ | 4 WHO IV (+7 WHO III) | Not specified. 1-3 per tumor | ✗ | ✓ | ✓ |
| Hu LS *et al.*; 2012 [8] | ✓ | 12 WHO IV (+12 other gliomas) | 21 | ✗ | ✓ | ✓ |
| Pathak AP *et al.*; 2001 [10] | ✗ | 12 rats with gliosarcoma | Over 3 slices per tumor | NA | ✓ | ✓ |
| Sadegui N *et al.*; 2008 [19] | ✓ | 2 WHO IV (+17 other gliomas) | 8 | ✗ | ✓ | ✓ |
| Sugahara T *et al.*; 1998 [20] | ✓ | 11 WHO IV (+19 other gliomas) | Not specified | ✗ | ✓ | ✗ |
| **Present study** | ✓ | 17 WHO IV | 71 | ✓ | ✓ | ✓ |

**Diagnostic MRI<sup>a</sup>:** Use of the initial MRI study performed as standard in the diagnosis of glioblastoma; **Spatial coregistration<sup>b</sup>:** Spatial coregistration between the MR images and the tissue samples; **Continuous histopathologic variable<sup>c</sup>:** Area/volume of microvascular structures; **NA:** Not applicable.

Two of the seven studies were developed with animal data [10, 17]; although the total number of subjects was higher in some human studies, the highest number of patients with glioblastoma was 12 patients in [8]; for all the studies, the number of tissue samples was low or not specified; none of the previous studies used the diagnostic presurgery MRI (took for diagnosis) to develop the correlation analyses; five of the seven studies used spatially correlated data and four of them analyzed continuous variables, as area or volume of vascular structures.

Considering those findings, our study seems to overcome most of these limitations. It includes data from a human cohort, with the highest number both of patients with glioblastoma and analyzed tissue samples. In addition, we uses the diagnostic MRIs, spatially corregistrated with histopathologic data and we analyze a continues histological variable. Furthermore, the method used to process MRIs is fully reproducible, automated and open-available.

### Statistical Analyses

#### Association between rCBV and MVA

Table 2 shows the descriptive measures of rCBV_mean_ and rCBV_max_ for the 71 blocks and the results of the Spearman correlation analyses between the MVP area and each imaging marker.

**Table 2:** Descriptive measures of rCBV_mean_ and rCBV_max_ (mean ± standard deviation, median and range) for the whole sample (71 blocks) and Spearman correlation results of these imaging markers with the microvascular proliferation area.

|  | **Mean ±**<br>**standard**<br>**deviation** | **Median** | **Range**<br>**[min, max]** | **Spearman correlation**<br>**with MVA**<br>**(Coefficient; pvalue)** |
| --- | --- | --- | --- | --- |
| $rCBV_{mean}$ | $4.92 \pm 2.69$ | 4.60 | [1.07, 13.42] | $C = 0.39$; $p = 0.0008^*$ |
| $rCBV_{max}$ | $9.57 \pm 5.01$ | 8.55 | [2.35 24.41] | $C = 0.42$; $p = 0.0002^*$ |

**MVA:** microvessel area

Both rCBV_mean_ and rCBV_max_ showed a significant positive correlation with MVA. That is, regions of the tumor with higher rCBV present significantly larger areas of microvessels.

Figure 4A shows an example (patient W33) of both rCBV and MVA maps to illustrate these two variables’ correlation. The white blocks did not present histopathological information, including the MVA data. It can be seen that blocks with areas with higher rCBV correspond with those with larger areas of MVA and vice versa.

**Figure 4.**
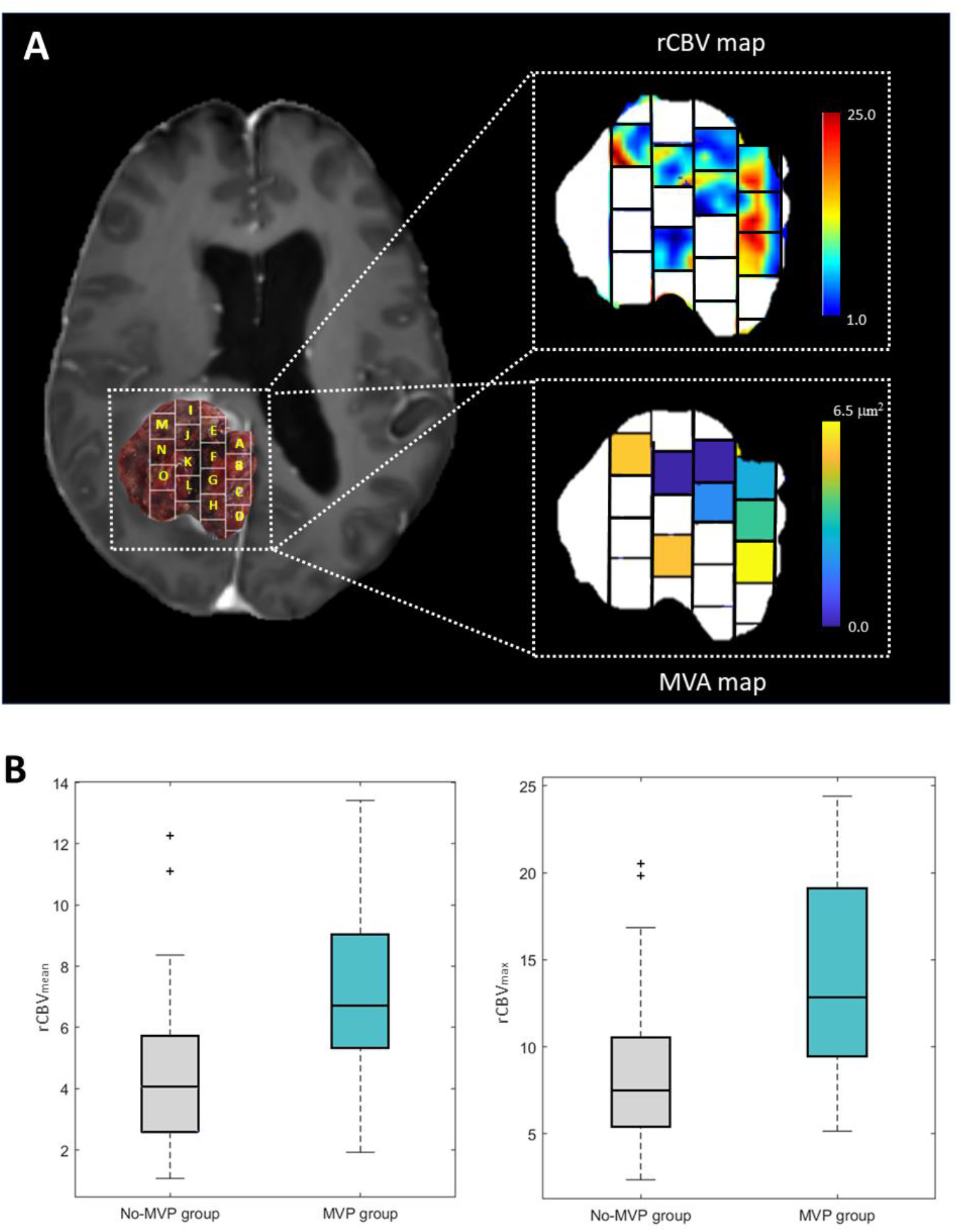
**A**: Post-gadolinium T1-weighted MRI overlaid with an image of the resected tumor including the delineation of the tissue blocks (left), example map of rCBV values (with a range of 1 to 25) of the areas occupied by the analyzed tumor blocks (top right), and example color map representing the MVA values (with a range from 0.0 to 6.5µ^2^) present in each analyzed block (below right). Blank blocks are not included due to lack of histopathological information. Example of patient W33 from the Ivy GAP database. **B**: Boxplot showing the significant differences of mean rCBV (left) and maximum rCBV (right) between *MVP group* and *non-MVP group*.

#### Differences of rCBV between MVP group and Non-MVP group

Table 3 shows the descriptive measures of rCBV_mean_ and rCBV_max_ for the blocks corresponding to the *MVP group* and the *Non-MVP group*, and the Mann-Whitney test results. All the measures (mean, minimum and maximum) of rCBV_mean_ and rCBV_max_ were higher in the *MVP group*, presenting values two times superior to the *non-MVP group*.

**Table 3:** Descriptive measures of rCBV_mean_ and rCBV_max_ (mean ± standard deviation and range) for the sample divided in the *MVP group* and *Non-MVP group* and Mann Whitney tests results from analyzing the differences of these imaging markers between these two groups are included.

|  | *MVP group*<br>(n = 14) |  | *Non-MVP group*<br>(n = 57) |  | MannWhitney<br>test |
| --- | --- | --- | --- | --- | --- |
| | Mean ± std | Range<br>[min, max] | Mean ± std | Range<br>[min, max] | p-value |
| $rCBV_{mean}$ | 7.08 ± 2.99 | [1.92, 13.91] | 4.38 ± 2.35 | [1.07, 12.25] | **p = 0.0006\*** |
| $rCBV_{max}$ | 14.15 ± 6.04 | [5.15, 24,41] | 8.44 ± 4.04 | [2.35 20.50] | **p = 0.0002\*** |

**MVP group:** microvascular proliferation group (including blocks with presence of microvascular proliferation); **Non-MVP group:** non microvascular proliferation group (including blocks with absence of microvascular proliferation); std: standard deviation; min: minimum; max: maximum.

The Mann-Whitney test yielded significant differences (p<0.05) between the rCBV_mean_ and the rCBV_max_ of the *MVP* and *non-MVP group*s. The rCBV_mean_ and rCBV_max_ can differentiate those blocks of the tumor with MVP from those blocks where there is not MVP.

Figure 4B shows the boxplots which illustrate these differences of the rCBV_mean_ and rCBV_max_ between the *non-MVP group* (lower values) and the *MVP group* (higher values).

## DISCUSSION

Microvascular proliferation is a histopathologic hallmark of glioblastoma, and MVA can be considered an independent prognostic biomarker according to previous studies’ results, in which authors reported significant longer survival in patients with glioblastoma tumors lacking MVP [10, 11]. Those works suggested that tumoral microvasculature is associated with survival differences among tumors with identical histologic grades [8, 10, 11]. However, despite its clinical potential value, direct MVA quantification is clinically unfeasible due to its time-consuming and labor-intensive nature.

By contrast, imaging markers derived from routinary MRI protocols, such as rCBV, present several advantages since they are fast to calculate, it does not represent any extra cost, and it is non-invasive compared with MVA. However, although rCBV is used for the assessment of brain tumors, it is not widely considered as a biomarker for clinical decision-making yet, probably due to the difficulty to normalize the rCBV values, which can generate confusion about prospective clinical guidelines, but also due to the lack of robust studies using spatially localized histologic correlations [16].

In this sense, here we investigated the association between the imaging markers rCBV_mean_ and rCBV_max_, calculated with a validated method [21-23], and the MVA in glioblastoma samples. Moreover, we analyzed the differences of rCBV between those areas of the tumor with MVP (with the presence of microvessels) from those regions of the tumor without microvascular proliferation (without microvessels).

We found significant correlations between rCBV_mean_ and rCBV_max_ and MVA when analyzing 71 tissue samples derived from 17 human glioblastoma tumors. Also, we found significant results when we evaluated the differences of both rCBV_mean_ and rCBV_max_ between tissue blocks with microvascular proliferation (*MVP-group*) from those blocks without MVP (*non-MVP group*).

Despite the few similar studies conducted [8-10, 16-20], our results are consistent with those previously reported, finding a significant positive correlation between rCBV and MVA and significant differences in rCBV in those regions of the tumor with MVP and those regions without MVP. In previous studies developed with human data and which analyze continuous variables (MVA or MV) [18, 19], similar correlation coefficients were found (ρ = 0.42 [18]; ρ = 0.46 [19] vs. ρ = 0.43 in the present study). However, in previous studies, only 2 and 4 glioblastoma patients were enrrolled, versus the 71 samples derived from 17 patients used in the current study. Increasing the interpatient heterogeneity in our study resulted in not higher correlation coefficients. Nonetheless, the analysis are more robust and p-values more significant.

We consider that this study provides a more solid evidence of the association between rCBV and MVA since a higher number of tissue samples of glioblastoma patients (n = 71) from the Ivy GAP public database was used, and since we used a method in which both the completely resected tumor and the corresponding magnetic resonance were spatially registered. Furthermore, we used an automated methodology [22, 23] that previously has been demonstrated in a multicenter study as robust when processing MRIs and calculating rCBV [21, 28], and that is publicly available in the ONCOhabitats site (www.oncohabitats.upv.es), ensuring the repeatability and reproducibility of the results.

However, this study has some limitations. Firstly, the manual registration between morphologic MRI images and the resected tumor image could be affected by deformations of the tumor tissue morphology when resected and/or the difficulty of finding matchings between both image features. Also, the number of independent analyzed samples is not much higher despite it being higher than in other previous studies.

The results derived from this work suggest the potential of imaging vascular markers calculated with the ONCOhabitats platform for addressing unmet challenges in glioblastoma management as a complementary tool of histologic markers in a clinical setting. We consider that the rCBV is a clinically relevant option for decision making in glioblastoma [16, 29], as it could be useful for analyzing intratumor vascular heterogeneity at both temporal and spatial levels in a non-invasive way [22].

The rCBV could be especially relevant as a diagnostic and prognostic marker for inoperable tumors, for which an exhaustive histopathological analysis cannot be performed. Furthermore, this study opens up the possibility of evaluating MVP more correctly after antiangiogenic treatments, in addition to other prognostic/predictive markers related to tumor vascularization. These results will be validated in a future prospective study with an independent patient cohort.

Concluding, the rCBV is shown as significantly different in those tissue blocks with MVP from those blocks without MVP, and it is significantly correlated with MVA, allowing spatial location and definition of different regions of the tumor since the first diagnostic stage in a non-invasive way.

## Data Availability

Data is available at https://doi.org/10.7937/K9/TCIA.2016.XLWAN6NL

